# Application of n-of-1 clinical trials in personalized nutrition research: a trial protocol for Westlake N-of-1 Trials for Macronutrient Intake (WE-MACNUTR)

**DOI:** 10.1101/2020.02.15.20023259

**Authors:** Yunyi Tian, Yue Ma, Yuanqing Fu, Ju-Sheng Zheng

**Affiliations:** Zhejiang Provincial Laboratory of Life Sciences and Biomedicine, Key Laboratory of Growth Regulation and Translation Research of Zhejiang Province, School of Life Sciences, Westlake University, Hangzhou, China; Institute of Basic Medical Sciences, Westlake Institute for Advanced Study, Hangzhou, China

**Keywords:** personalized nutrition, dietary intervention, n-of-1, single patient trial, high-fat diet, low carbohydrate diet, postprandial blood glucose, gut microbiome

## Abstract

Personalized dietary recommendations can help with more effective disease prevention. This study aims to investigate the individual postprandial glucose response to diets with diverse macronutrient proportions at both individual level and population level and explore the potential of the novel single-patient (n-of-1) trial for the personalization of diet. Secondary outcomes include individual phenotypic response and the effects of dietary ingredients on the composition and structure of gut microbiota. Westlake N-of-1 Trials for Macronutrient Intake (WE-MACNUTR) is a multiple crossover feeding trial consisting of three successive 12-day dietary intervention pairs including a 6-day wash-out period before each 6-day isocaloric dietary intervention (a 6-day high-fat, low-carbohydrate (HF-LC) diet and a 6-day low-fat, high-carbohydrate (LF-HC) diet). The results will help provide personalized dietary recommendation on macronutrients in terms of postprandial blood glucose response. Well-designed n-of-1 trial is likely to become an effective method of optimizing individual health and advancing health care. This trial is registered at clinicaltrials.gov (NCT04125602).

## Introduction

Diet and nutrition are key to maintain human health. Previous studies have showed much interest in the metabolic effects of different ratios of dietary fat to carbohydrate intake(1-5). Some evidence suggests that a high-fat, low-carbohydrate (HF-LC) diet can improve glycemic control by reducing glycated hemoglobin and fasting glucose, while others support the beneficial effect of a low-fat, high-carbohydrate (LF-HC) diet with particular focus on the quality of carbohydrate(1, 6-10). One important interpretation for the inconsistent results is the individualized or personalized response to the dietary macronutrient intake, which is also called “personalized nutrition”(11).

Personalized nutrition focuses on an individual’s potentially unique dietary needs instead of assuming a “one-size-fits-all” approach where everyone is thought to benefit from the same diet (12). The general aim of personalized nutrition is to improve health using nutritional, genetic, phenotypic and other information of the individuals to develop targeted nutritional advice, services or other products(13-15). Although specific dietary recommendation has been made for pregnant women, infants, children, adults or elderly, they are still subgroup recommendation, which is far from the stage of “personalization” or “precision”.

Application of ‘n-of-1’ clinical trial or ‘single patient’ study represents a new direction in personalized nutrition research (**Figure 1**). It can capture intra-individual variability in health behaviors over time, aiming to identify individual response to a given intervention in a controlled trial, which provides great opportunity to answer the personalization potential of different diets, nutrients or nutrition supplements(16-18). The idea of n-of-1 has been applied in special education, psychotherapy, psychology and pharmaceutical studies for decades to test the individual response to specific drugs or treatments(19-23). Stunnenberg et al. reported the efficacy of mexiletine on reducing muscle stiffness in patients with nondystrophic myotonia (NDM) using a series of n-of-1 trials(24). However, there is no published ‘n-of-1’ study in nutrition field so far. Westlake N-of-1 Trials for Macronutrient Intake (WE-MACNUTR) is a novel series of clinical trials that use the macronutrient intake as an exemplar for the new progress in the field.

**Figure 1.**
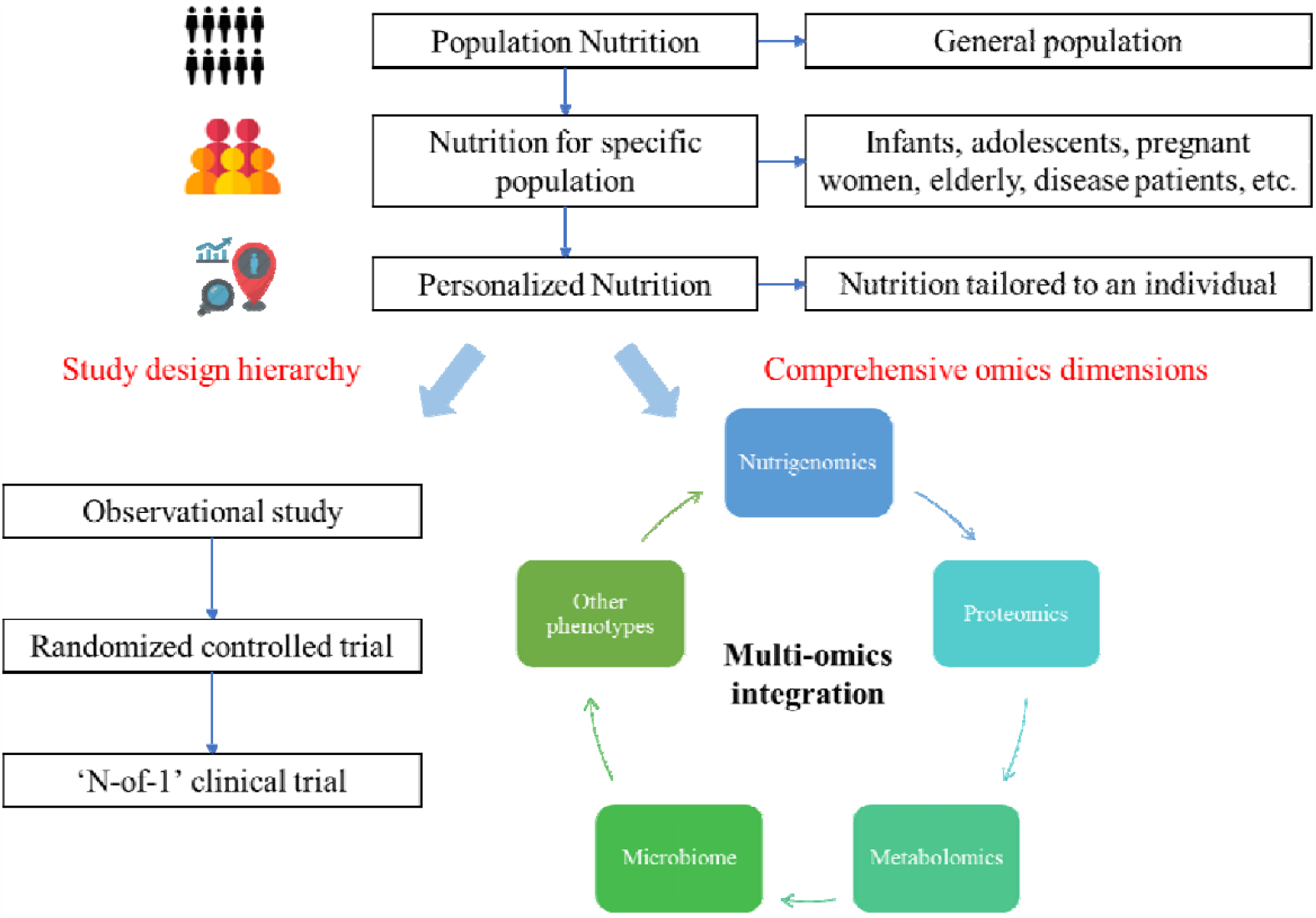
The Development of Personalized Nutrition. Personalized nutrition was born in the context that a conventional ‘one-size-fits-all’ approach usually fail to meet individual’s need for the nutritional requirements. “N-of-1” clinical trial is a novel study design for the research of personalized nutrition comparing with the traditional designs such as observational study and randomized controlled trial. Integration of multi-omics data including nutrigenomics, proteomics, metabolomics, microbiome and other phenotypes is key for the development of personalized nutrition.

We will describe the trial protocol of the WE-MACNUTR: a series of n-of-1 feeding clinical trials among healthy adults. The feeding trial is composed of two dietary interventions, a HF-LC diet and a LF-HC diet. The primary objective is to investigate the response of postprandial blood glucose to the dietary interventions and the primary outcomes include postprandial maximum glucose (PMG), the area under the curve (AUC24) of postprandial glucose from 0.00 to 24.00 and the mean amplitude of glycemic excursions (MAGE) obtained by measuring the arithmetic mean of the differences between consecutive peaks and nadirs. Secondary objectives of this study include evaluating different phenotypic responses, such as circulating lipid profile changes to a specific diet among individuals and evaluating the impact of different dietary components on the composition and structure of gut microbiota.

## Methods

### Study Design

The present n-of-1 trial is a multiple crossover feeding trial conducted in a single participant, comparing her/his response to different interventions and assessing the variability in these responses(25). In the WE-MACNUTR study, a series of n-of-1 trials are employed simultaneously and a common regimen of interventions is applied to all participants (**Figure 2**). Participants experience three successive 12-day intervention pairs including a 6-day wash-out period between each intervention. The diets are isocaloric and all provided by the researchers, and the primary distinguishing feature is their fat and carbohydrate contents, including a HF-LC diet and a LF-HC diet. Prior studies have shown that dietary pattern may have a rapid influence on glycemic control. Parr and colleagues performed an intervention study and successfully observed a significant effect of high-carbohydrate diet (compared with a high-fat diet) on various blood glucose measurements after a 5-day intervention(26). In addition, gut microbial community was also reported to change substantially in response to the dietary change within four days(27). Therefore, we set a wash-out period of six days between each intervention arm (HF-LC and LF-HC), after balancing the outcomes of the study (postprandial glycemic response, gut microbiota change), and feasibility of the present feeding trials. Each 12-day intervention pair comprises 6 days of HF-LC (three meals daily) and 6 days of LF-HC (three meals daily) diets in a random order. The order of the diets in each pair is determined using a block randomization. Major investigators and laboratory personnel responsible for the measurements are masked to group allocation. Meal providers are aware of group and diet allocation, but they are not involved in the rest of the trial.

**Figure 2.**
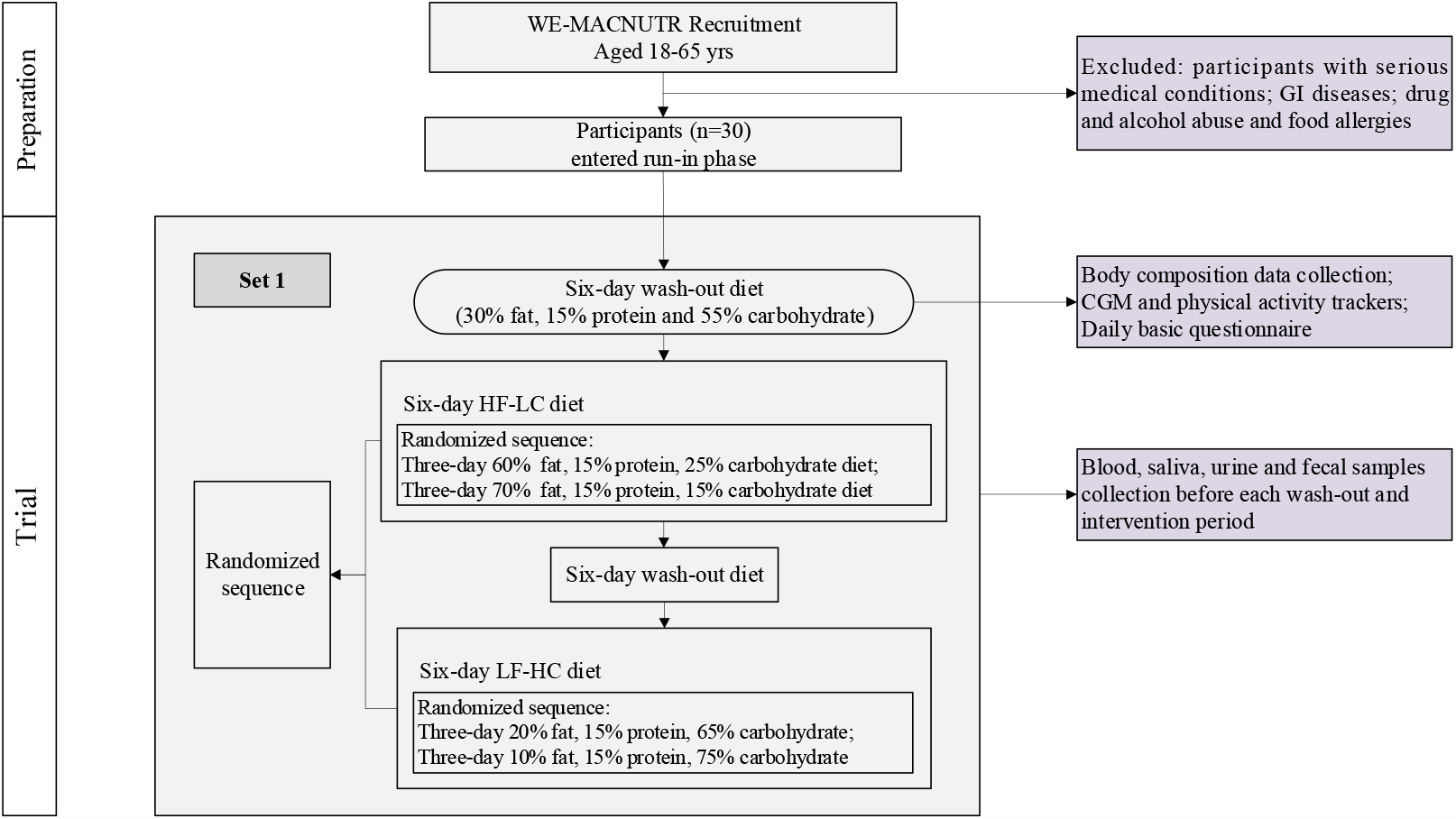
Flow Diagram of the Westlake N-of-1 Trials for Macronutrient Intake (WE-MACNUTR) Trial. The flowchart summarizes the preparation phase and the first set of the trial. Set 2 and set 3 share the same trial design as set 1 except for no blood sample collection before each wash-out or intervention period. The sequence of two types of six-day dietary interventions in each set is randomized using block randomization as LF-HC and HF-LC diets in set 1; HF-LC and LF-HC diets in set 2 and HF-LC and LF-HC diets in set 3, respectively. CGM: continuous glucose monitoring; GI diseases: gastrointestinal diseases; HF-LC: high-fat, low-carbohydrate; LF-HC: low-fat, high-carbohydrate; WE-MACNUTR: Westlake N-of-1 Trials for Macronutrient Intake.

Participants are required to complete a basic questionnaire on a daily basis after dinner to summarize their eating behaviors and multiple lifestyle factors including physical activity, mood and sleep patterns throughout the day. Biological samples including blood, saliva, urine and fecal samples will be collected every 6 days for metabolomics profiling using untargeted platforms. Saliva will also be collected for oral microbiota investigation (**Figure 3**). The fasting venous blood samples will be collected every 6 days only during the first set of the study as to reduce the burden and increase the compliance of the participants. The participants will be asked to wear a continuous glucose monitoring (CGM, Freestyle Libre Pro System), which measures interstitial glucose every 15 minutes. The same CGM system was used by a recent personalized nutrition study (PREDICT 1) to monitor individual postprandial glycemic responses(28). As most individuals do not resemble “the average” in the context of precision nutrition, using data sets generated from digital wearable devices, such as CGM, will potentially help inform individualized food choices(29). In the present study, participants will be asked to wear the sensor two days before the start of each intervention period, and remove it by the end of each 6-day intervention period. CGM will not be used during the washout periods. In addition, participants are asked to continue their regular daily activities and exercise throughout the study period, and wear a wrist-based triaxial accelerometer (AX3, Axivity) on the non-dominant wrist to monitor the physical activity and exercise intensity during the intervention period.

**Figure 3.**
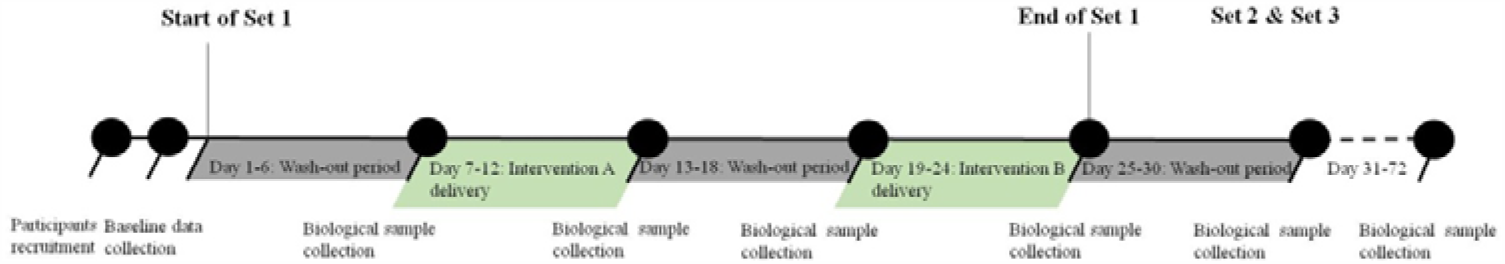
The Timeline of the Westlake N-of-1 Trials for Macronutrient Intake (WE-MACNUTR) Trial. The timeline illustrates a preparation period for participants recruitment, a baseline data collection period, and three feeding trial periods. The first set of the trial consists of two wash-out periods (highlighted in green) and two randomized dietary intervention periods (highlighted in grey). In all three sets of the trial, both wash-out and intervention periods last for 6 days respectively.

The study was approved by the Westlake University Internal Ethical Review Board in Hangzhou, China, and registered with ClinicalTrials.gov (identifier: NCT04125602). Written informed consent are obtained from all study participants.

### Dietary Intervention

A dietitian designs the diet for the intervention and the washout period based on the Chinese Dietary Guidelines (2016) and Chinese Dietary Reference Intakes (2013) as well as the participants’ demographic information, eating habits and physical activity levels(30, 31). Besides, factors such as local food availability, the cooking methods of the Westlake University canteen kitchen, and the current recipes are important factors in the meal planning. Participants are instructed to consume only the provided foods or beverages.

#### Wash-out Diet (WD)

Prior to each dietary intervention, all participants are provided with the same standardized diets for 6 days as the ‘wash-out diet’ to reduce potential sources of bias and eliminate any carry-over effect of previous intervention. The wash-out diet consists of 30% of total energy (%E) from fat, 15%E from protein and 55%E from carbohydrate based on the acceptable macronutrient distribution range (AMDR)(32).

#### HF-LC Diet Intervention

Throughout the 6-day HF-LC intervention, participants are provided with a HF-LC diet, including a three-day diet consisting of 60%E from fat, 15%E from protein and 25%E from carbohydrate while the other three-day diet consisting of 70%E from fat, 15%E from protein and 15%E from carbohydrate.

#### LF-HC Diet Intervention

Throughout the 6-day LF-HC intervention, participants are provided with a LF-HC diet, including a 3-day diet consisting of 20%E from fat, 15%E from protein and 65%E from carbohydrate while the other 3-day diet consisting of 10%E from fat, 15%E from protein and 75%E from carbohydrate.

### Participants

Participants are recruited among students and staffs from Westlake University, Hangzhou, China. Inclusion criteria are: adults aged between 18 and 65 years; participants are able to provide written informed consent, and have access to smart phones or computers. Exclusion criteria include: long-term gastrointestinal diseases; neurological conditions and cognitive impairment; other clinically diagnosed medical conditions including type 2 diabetes, hypertension, cardiovascular diseases, liver, kidney diseases and/or other systemic diseases; taking antibiotics within the last two weeks; hospitalization or surgery planned within the next three months; pregnant or lactating women; tobacco, alcohol, or illicit drug abuse; vegan or food allergies; with no access to a smart phone or computer with an internet connection; enrolled in concurrent intervention study; and non-Chinese speaking participants. This study emphasizes more on assessing individual responses to the intervention of interest rather than drawing a general conclusion at population level, and therefore, study participants cover a broad age range and will not be balanced by sex.

### Measures

The primary outcomes are (1) postprandial maximum glucose (PMG). PMG is the peak value of CGM within 3 hours after the first bite of a meal or the maximum value of CGM between two meals when the interval is less than 3 hours. (2) area under the curve (AUC24). AUC24 refers to total area under the CGM curve from 0.00 to 24.00. (3) mean amplitude of glycemic excursions (MAGE). MAGE is obtained by measuring the arithmetic mean of the differences between consecutive peaks and nadirs, provided that the differences are greater than 1 SD around the mean glucose values. Secondary outcomes include different phenotypic responses, such as circulating lipid profile and gut microbiome profile, to a specific diet among individuals (**Table 1**).

**Table 1.** Outcome and Assessment Points.

| Variable | Measure | Assessment point |  |  |
| --- | --- | --- | --- | --- |
|  |  | T<sub>p</sub>S1<sup>a</sup> | T<sub>p</sub>S2<sup>b</sup> | T<sub>p</sub>S3<sup>c</sup> |
| Primary outcomes |  |  |  |  |
| Continuous glucose level |  |  |  |  |
| PMG |  | ● | ● | ● |
| AUC24 | FreeStyle Libre Flash | ● | ● | ● |
|  | Glucose Monitoring System |  |  |  |
| MAGE |  | ● | ● | ● |
| Secondary outcomes |  |  |  |  |
| Lipid metabolism | Blood samples |  |  |  |
| Inflammation | Blood samples |  |  |  |
| Oral microbiota | Saliva | √ | √ | √ |
| Gut microbiota profiling | Fecal samples | √ | √ | √ |
| Fecal metabolites | Fecal samples | √ | √ | √ |
| Metabolomics profiling | Blood samples |  |  |  |
|  | Fecal samples | √ | √ | √ |
|  | Urine samples | √ | √ | √ |
| Physiological characteristics |  |  |  |  |
| Weight | Kubei height scale |  |  | √ |
| Blood pressure | YUWELL YE660D upper arm sphygmomanometer |  |  | √ |

Abbreviations: AUC24, total area under the continues glucose monitoring curve from 0.00 to 24.00; MAGE, mean amplitude of glycemic excursions; PMG, postprandial maximum glucose.
<sup>a</sup>T<sub>p</sub>S1 represents set 1.
<sup>b</sup>T<sub>p</sub>S2 represents set 2.
<sup>c</sup>T<sub>p</sub>S3 represents set 3.
● per day during intervention periods
√ before and after intervention periods

### Sample Size Calculation

In the present study, a total of 30 participants will be enrolled. Bayesian hierarchical model meta-analysis will be applied to combine the results from each n-of-1 trial to generate pooled effect estimate at population level. However, no formula-based methodology exists for sample size calculation for such design at population level(33). Referring to the method reported by Stunnenberg et al., we performed a simulation-based statistical power calculation at population level with modifications(24). In brief, a prior distribution for the mean intervention effect (HF-LC vs. LF-HC) on the PMG was prespecified based on results of a previous RCT reported by Parr et al(26). In their study, 1.6 mmol/L (6.2 mmol/L vs. 7.8 mmol/L) difference in the PMG was established between participants receiving high-fat versus high-carbohydrate meals. We drew a random realization and simulated an individual-specific virtual mean intervention effect for each participant. At the next level of simulation, longitudinal measurements for each participant (3 sets, 2 intervention periods per set and 18 observations per intervention period) were simulated assuming a normal distribution of the measurements centered around the individual-specific virtual mean effect size, with a common within-participant residual variance. Thereafter, all the simulated data from these n-of-1 trials were used to perform a multilevel Bayesian meta-analysis specifying a linear mixed model with flat non-informative priors at population level.

A previous study found that the difference in PMG between healthy participants aged 25-45 years and those older than 45 years was 3mg/dL (0.167mmol/L)(34, 35). Therefore, we considered this magnitude of difference to be clinically meaningful and determined the posterior probability on both intervention effect of at least 0.167 mmol/L from the simulation-based Bayesian meta-analysis based on the simulation data. In order to keep a balance between the power of detecting meaningful differences and false positive rate, a relatively strict 90% posterior probability on an intervention effect of 0.167 was treated as a clinically positive result at population level, while a stricter 99% posterior probability on an intervention effect of not being equal to zero was treated as a statistically positive result. We repeated the above procedures for 1000 times (corresponding with data from 1000 aggregated N-of-1 trials) and the fraction of meta-analyses that returned a positive conclusion was treated as measure for the power. Consequently, the power was estimated to be 100% for detecting the intervention effect of at 0.167 mmol/L difference in PMG.

On the other hand, in order to determine the false positive rate, we performed another set of simulation under the assumption of no intervention effect. For each virtual participant, we simulated data that allowed the participant to have an individual-specific virtual mean intervention effect which centered around the true population-level mean effect size (zero) with a normal distribution. At the next level of simulation, longitudinal measurements for each participant (3 sets, 2 intervention periods per set and 18 observations per intervention period) were simulated assuming a normal distribution of the measurements centered around the individual-specific virtual mean effect size, with a common within-participant residual variance. All data from these 30 virtual individual N-of-1 trials were used to perform the multilevel Bayesian meta-analysis at population level. A posterior probability of more than 90% on an intervention effect of 0.167 mmol/L and more than 99% on an intervention effect of not being equal to zero were treated as clinically and statistically false positive results, respectively. After repeating the procedures for 1000 times, the fraction of meta-analyses that returned a clinically or statistically false positive conclusion was determined as measure for the type I error rate at population level, which was found to be 0.023 and 0.048, respectively. Thus, this simulation-based sample calculation indicates that with 30 participants completing the trial (3 sets, 2 intervention periods per set and 18 observations per intervention period), we will have satisfactory type I error rate and enough power to detect the prespecified intervention effect.

## Statistical Analysis Plan

### Data Management

Even though all participants are students/staffs who routinely have meals in campus, we anticipate that some participants will skip some meals that we provide at dining room and eat other foods instead, which adds uncertainty to the effects on postprandial blood glucose. On this occasion, we will ask the volunteers to record all the other foods they have eaten not provided by the researchers. Other major violations, such as failure to complete at least one set of intervention, will prevent statistical analysis at individual level and lead to exclusion of the participants from meta-analysis at population level.

### Primary Analysis

The primary analysis of intervention effect is the comparison of the effect of HF-LC diet with that of LF-HC diet on postprandial blood glucose level. At individual level, we use individual intervention effects to guide dietary decisions for each participant. On the other hand, we generate estimates of intervention effect at population level by combing the n-of-1 results with meta-analysis.

### Analysis of Baseline Data

Descriptive statistics with demographics and baseline characteristics will be presented for each participant.

### Analysis of Individual N-of-1 Trials

Statistical analysis will be performed separately for each n-of-1 trial to estimate the intervention effect at individual level. Bayesian models will be applied to estimate the intervention effects (**Figure 4**). Posterior probabilities of outcomes will be calculated using an interface that incorporates open-source R software (3.6.1) and open-source OpenBUGS (version 3.2.3). The results will be reported for primary variables (e.g., peak concentration of postprandial blood glucose and AUC of postprandial glucose concentration over 24 hours), and participants will be provided with an estimate of differences in the variables and the probabilities that the differences are induced by the different interventions (HF-LC vs. LF-HC).

**Figure 4.**
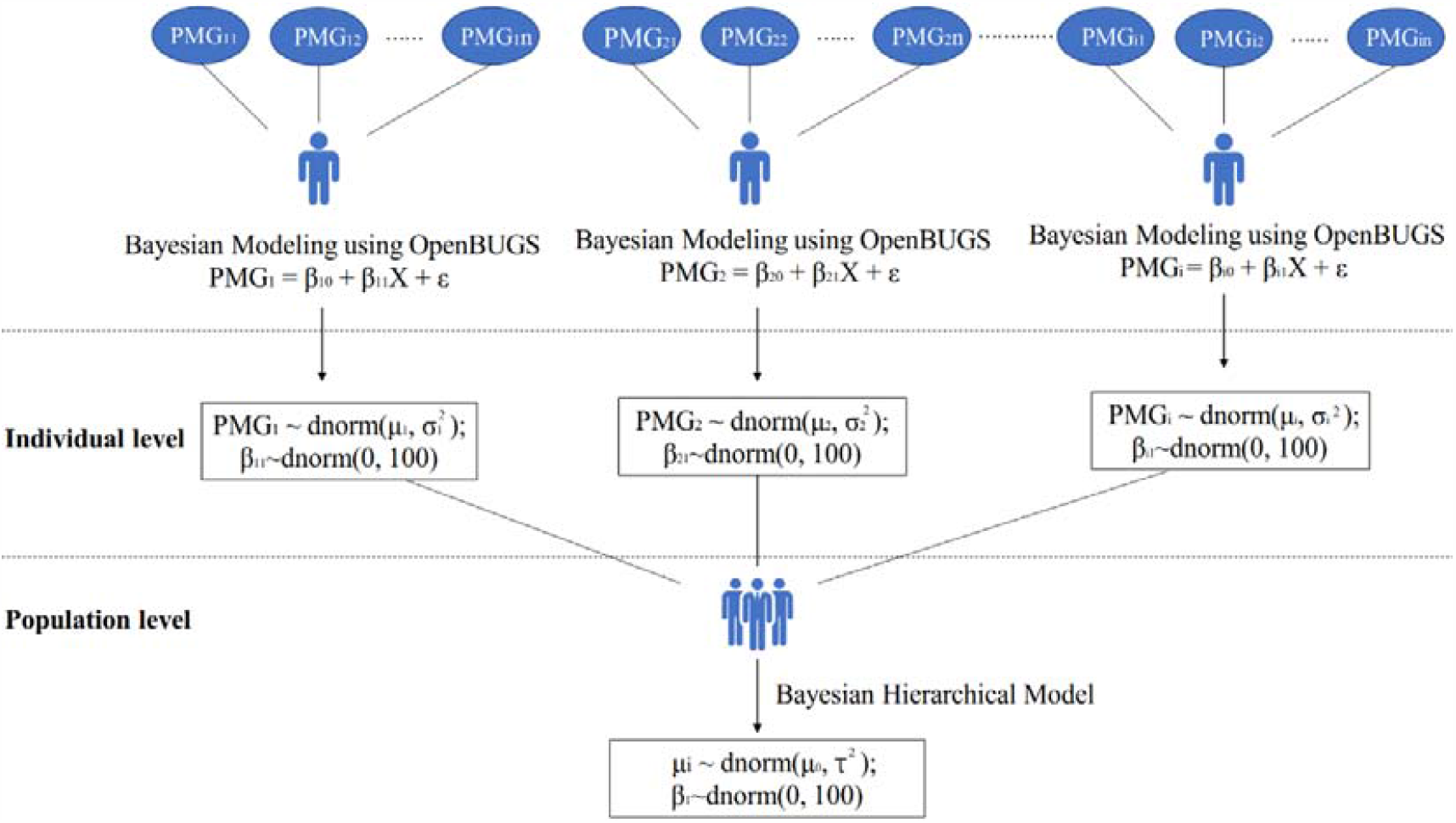
Representation of the Hierarchical Bayesian Estimation for the Primary Outcomes at Both Individual and Population Level (combination of single patient studies). The observed repeated measurements of the postprandial peak glucose on a given patient are combined into a sample mean and sample variance. The model assumes that the patients’ measurements follow a normal distribution centered about that patient’s true mean effect (μ_i_) with variance σ_1_^2^. At the population level, the various patients’ true mean (μ_i_) are assumed to follow a normal distribution centered about an overall population mean (μ_0_) with between-patient variance τ^2^. For the Bayesian specification, prior distributions are assigned for β, μ_0_, σ_1_^2^, and τ^2^. In the present study, these prior distributions are standard non-informative prior distributions. X means the independent variable: dietary pattern (high-fat and low-carbohydrate vs. low-fat and high-carbohydrate). Secondary and exploratory outcomes will be analyzed similarly. PMG: postprandial maximum glucose.

### Meta-analysis of N-of-1 Trials

A Bayesian multi-level model will be used to combine results of the multiple n-of-1 trials(24, 36, 37). Participant will be treated as a random effect and a common within-participant residual variance will be assumed. Non-informative priors will be applied for all model parameters, with mean parameters using normal prior distributions with very large standard deviations and variation parameters using inverse gamma distributions with both shape and scale parameters equal to 0.01. Using the interface that incorporates R software (3.6.1) and OpenBUGS (version 3.2.3), combining the data from the individual n-of-1 trials will obtain posterior distributions for the mean intervention effect at the population level. Secondary and exploratory outcomes will be analyzed similarly.

## Discussion

To advance the field of personalized nutrition, n-of-1 clinical trial appears to be a promising study design to advocate, although the real-world example is rare. We will use the WE-MACNUTR trial as an exemplar to showcase the study design of n-of-1 trial as to test the individualized response to macronutrient intake among healthy adults. The study, if successful, will provide insight into the feasibility of n-of-1 approach to personalize or tailor dietary intervention to individuals.

Previous study suggested that the magnitude of postprandial responses to mixed meals depended largely on the total amount of fat and carbohydrates intakes(1, 5, 6). The American Diabetes Association (ADA) recommends monitoring carbohydrate intake to achieve better glycemic control in patients with type 2 diabetes, which is based on studies showing reduced postprandial glucose and triglyceride responses in individuals consuming HF-LC diets(38-40). However, previous systematic reviews discussed the effects of HF-LC and LF-HC diets on metabolic risk factors and showed inconsistent results(5, 7, 41). Hsu et al suggested LF-HC diet consisting of high fiber contents showed beneficial effects on glucose and insulin sensitivity among both Asian Americans and Caucasian Americans(42). Several studies have reported that both HF-LC and LF-HC diets reduced HbA1c and fasting glucose in obese adults with T2D, while HF-LC diet achieved better improvements in glycemic control(1, 2). Kamada et al suggested that changes in HbA1c and fasting plasma glucose did not differ significantly between HF-LC and LF-HC diets among Japanese diabetic patients(43). The benefits and drawbacks of different dietary patterns on health are under intensive study these days despite the lack of a standardized definition regarding the macronutrient contents(44-46). Therefore, n-of-1 trial has a huge potential to help explore the main effects of a specific dietary intervention, and identify the factors that influence individual response to nutritional factors. In the present study, it is expected that the trial will provide information on the response of postprandial blood glucose level among individuals to different dietary interventions, namely HF-LC and LF-HC diets, enabling better understanding of intra-individual differences in absorption, distribution and metabolism of macronutrients.

Individual human beings are not only unique in the aspect of host genome, but also in the aspect of the gut microbiome, which represents the combined influence of the diet and lifestyles, as well as host genetics(47, 48). Both animal and human studies have demonstrated that the composition of the gut microbiome can be rapidly affected by a specific dietary component exposure within four days(27, 49). Furthermore, integration of machine learning algorithms with gut microbiome features have shown powerful potentials to predict one’s response to different dietary patterns in term of postprandial glucose response(50). Zeevi *et al*. monitored the postprandial glucose response in a cohort of 800 participants in Israel in response to identical meals. A multidimensional data including gut microbiome features, anthropometrics, blood parameters, and physical activity were integrated into a machine learning algorithm that was capable of predicting personalized postprandial glucose response with the gut microbiota(51). These new progresses have stimulated more research on the application and integration of gut microbiome into the personalized nutrition field. Results from this study may provide further evidence suggesting that n-of-1 trial is feasible in characterizing individual microbiome profile.

With the aggregated data from isocaloric meal but different carbohydrate to fat ratio, our study will make deeper investigations of the underlying interactions between specific food components and microbiota species. Therefore, future methodological studies on developing and implementing effective evaluation of personalized dietary intervention could assist individuals in promoting healthy gut microbiota profile and preventing cardiometabolic diseases. Another strength of the present study with n-of-1 method is its flexibility which enables the study design to be personalized to individuals’ interests and requirements, and its high level of evidence for making clinical decisions for individuals alongside systematic reviews of RCTs.

## Limitations

The n-of-1 study does have limitations. Participation in feeding trial such as WE-MACNUTR requires time and effort so the trial cannot be conducted in an ideally controlled setting. Compliance of the participants to the intervention over time is challenging, as they are all required to eat the provided foods with no extra food intake throughout the feeding trial. Any extra snack or beverage intake could affect individual blood glucose level. Besides, slight changes in cooking method, food groups or food ingredients under inevitable circumstances would also jeopardize the final results.

## Conclusions

In summary, WE-MACNUTR trial, as an exemplar of nutritional n-of-1 trial, will address the call for new method to advance the field of personalized nutrition. WE-MACNUTR will potentially help clarify the individual postprandial glucose response to diets with diverse macronutrient proportions, and help design and optimize macronutrient composition in long-term dietary intervention studies. The results of WE-MACNUTR will also be helpful in terms of understanding the individual response of gut microbiome to macronutrients.

## Data Availability

The corresponding author will provide access to individual de-identified participant data from the present study, upon request by qualified researchers whose proposals are approved. Requests for access to the individual-level data from this study can be submitted via email to and proposals are required to be attached for approval.

## Abbreviations used

AUC24: area under the curve
CGM: continuous glucose monitoring
HF-LC: high-fat, low-carbohydrate
LF-HC: low-fat, high-carbohydrate
MAGE: mean amplitude of glycemic excursions
PMG: postprandial maximum glucose
WE-MACNUTR: Westlake N-of-1 Trials for Macronutrient Intake.

## Acknowledgements

We thank all the volunteers participating in the Westlake N-of-1 Trials for Macronutrient Intake (WE-MACNUTR). The authors’ contributions were as follows: JSZ designed the study and was the principal investigator of WE-MACNUTR; YT, YM, YF and JSZ drafted the manuscript; and all authors read and approved the final manuscript.

## Notes

### Competing Interest Statement

The authors have declared no competing interest.

### Clinical Trial

NCT04125602

### Funding Statement

This study was funded by National Natural Science Foundation of China (81903316), Zhejiang Ten-thousand Talents Program (2019R52039) and Westlake University (101396021801).

